# Chinese Translation and Psychometric Testing of the Peripheral Intravenous Catheter Insertion Self-Confidence Scale

**DOI:** 10.1101/2025.11.25.25341024

**Authors:** Zhenzhen He, Pei Yu, Xuejing Jia, Lianlian Hu, Zhijie Xue, YiQing Lü, Caroline Marchionni

## Abstract

**Background and Purpose:** There is no tool in China to evaluate self-confidence in peripheral intravenous catheter insertion,this study aims to translate the Peripheral intravenous Catheter Insertion Self-Confidence Scale (PVCS) into Chinese and evaluate its reliability and validity among nursing students.

**Methods:** The translation and validation followed Brislin’s model, including forward translation, back-translation, synthesis, expert review, and pre-testing to develop the Chinese version (C-PVCS). A convenience sample of 205 nursing students from Tianjin and Chongqing, China, was surveyed to assess reliability and validity

**Results:** The Chinese version of the PVCS (C-PVCS)comprises 15 items in two dimensions: theoretical knowledge self-confidence (5 items) and operational skills self-confidence (10 items). The scale’s Cronbach’s α coefficient was 0.979, the split-half reliability was 0.944, and the test-retest reliability was 0.985. The average scale-level content validity index (S-CVI/Ave) was 0.966, and the item-level content validity index (I-CVI) ranged from 0.875 to 1.000. Exploratory factor analysis extracted two common factors, with a cumulative variance explained of 85.839%. For confirmatory factor analysis, the structural equation model fitting indices showed that the root mean square error of approximation (RMSEA) was 0.077, and the chi-square/degree of freedom ratio (χ2/df) was 2.22.

**Conclusion:** The C-PVCS demonstrates excellent reliability and validity, making it a suitable tool for assessing nursing students’ self-confidence in peripheral intravenous catheter insertion.

## Introduction

Peripheral intravenous catheter insertion is among the most common invasive procedures in clinical practice(LI,L et al, 2024). It is often used for short-term fluid and drug administration, and approximately 75%-95% of hospitalized patients require intravenous fluid therapy(Jacobs, 2022; Keleekai et al, 2016; LI,L. et al, 2024). In this context,self-efficacy refers to an individual’s belief in their ability to successfully complete a specific behavior or task(Cruz et al, 2023). whereas “self-confidence” refers to the belief that one has the ability to successfully perform peripheral intravenous catheter insertion. Research has shown a direct correlation between the level of self-confidence in peripheral intravenous catheter insertion and the first-attempt success rate(Keleekai et al, 2016). Catheter insertion failure not only causes fear, pain, and decreased satisfaction for the patient, but also leads to complications like hematoma and nerve damage, while increasing the rate of infection and delaying medication time, which in turn increases hospital costs(Marsh et al, 2021). Currently, there is no tool in China to evaluate self-confidence in peripheral intravenous catheter insertion. In 2023, Marchionni et al(Marchionni et al, 2024)developed the PVCS by revising and combining instruments from Schuster et al(Schuster et al, 2016) and Engum et al(Engum et al, 2003). The scale has since been translated and implemented in Turkey in 2024(Özbay et al, 2025). This study aims to translate this scale into Chinese and test its reliability and validity, providing a reliable tool for assessing the self-confidence level of nursing students in peripheral intravenous catheter insertion.

## Methods

### 1.1. Description of the Original Peripheral intravenous Catheter Insertion Self-Confidence Scale

The original scale has three dimensions and 15 items. These are “learning self-confidence” (4 items), “preparation and securement self-confidence” (4 items), and “insertion self-confidence” (7 items). A 5-point Likert scale is used to evaluate the content, with options ranging from “strongly disagree” to “strongly agree”, corresponding to scores of 1 to 5, respectively. The scores for all items are summed to get a total score ranging from 15 to 75,a higher total score indicates greater self-confidence in peripheral intravenous catheter insertion. The overall Cronbach’s α coefficient of the scale is 0.89, with dimension αs ranging from 0.72 to 0.85, demonstrating good reliability.

### 1.2. Scale Translation and Cultural Adaptation

#### 1.2.1. Scale Translation

After obtaining email permission from the scale’s author, the translation was conducted following the Brislin translation model(GUO,J.Y. & LI,Z., 2012). ① Forward translation: One nursing master’s student with a CET-6 English level and one university English major teacher independently translated the original scale to create versions T-1 and T-2. ② Synthesis: A university nursing teacher systematically compared the original scale with T-1 and T-2 to produce a synthesized T-3 version. Three translators then discussed T-1, T-2, and T-3 to reach a consensus, forming the official forward-translated version T. ③ Back-translation: Two Ph.D. students in nursing separately translated the T version back into English, forming BT-1 and BT-2. A master’s student majoring in English then compared T with BT-1 and BT-2. After an in-depth discussion, a back-translated version, BT-3, was finalized. ④ Expert Committee Review: A committee consisting of three forward translators and three back-translators discussed and compared the original scale, forward-translated versions (T-1, T-2, T-3), and back-translated versions (BT-1, BT-2, BT-3). They revised any inappropriate or ambiguous expressions and, after reaching a consensus, selected the most appropriate wording to create the final back-translated version BT. ⑤ Original Author Review: The forward-translated T version and the back-translated BT version were sent to the original author for feedback and correction. After receiving a unified opinion, the Chinese version of the Peripheral intravenous Catheter Insertion Self-Confidence Scale was finalized.

#### 1.2.2. Cultural Adaptation and Content Validity Check

This study invited a group of eight experts with extensive academic backgrounds in nursing or education and long-term experience in nursing management to form an expert panel. They were tasked with conducting a cross-cultural adaptation and content validity evaluation of the scale. The expert panel included two deputy chief nurses, one chief physician in nephrology, two nursing education experts, and three clinical nurse managers. The panel consisted of two males and six females, with an average of 24-34 (25.87 ± 5.13) years of work experience. One expert held a doctoral degree, four held master’s degrees, and three held bachelor’s degrees. Two experts had senior professional titles, and three had vice-senior professional titles. The experts were asked to systematically evaluate the scale based on their work experience and expertise, focusing on the relevance of the scale items and the clarity of their expression. During the expert group review, special attention was paid to the cultural adaptability of the items. Some vocabulary and contextual expressions were adjusted to ensure they align with the cognitive habits and linguistic preferences of Chinese nursing students. Based on these revisions, the researchers further refined the scale, resulting in the C-PVCS.

#### 1.2.3. Pre-survey

This study conveniently selected 20 trainee nurses from a hospital in Tianjin to participate in a pre-survey using the Chinese version of the Peripheral intravenous Catheter Insertion Self-Confidence Scale. The inclusion criteria were: (1) currently enrolled nursing students who had systematically studied the peripheral intravenous catheter insertion course, and (2) had normal comprehension and communication skills and voluntarily participated in the study. The exclusion criterion was: nursing students who had not systematically studied the peripheral intravenous catheter insertion course. Before the study, the nurses were informed of the purpose and significance of the study and asked to scan a QR code via WeChat to complete the survey. During the survey, they were asked if they understood the scale’s meaning and if there was any ambiguity in the content, and their questions were answered immediately. The time required to complete the questionnaire was also recorded. Based on the pre-survey results, the final C-PVCS was determined.

### 1.3. Reliability and Validity Testing

#### 1.3.1. Participants

The study participants were nursing students from three universities in Tianjin and Chongqing. The sample size calculation is based on the number of items in the scale, set at 5-10 times the number of scale items(WEI,Y.T. et al, 2025),plus 20% attrition (e.g., due to missing data and invalid questionnaires), targeting 94-188 samples (actual n=205). The inclusion and exclusion criteria were the same as those for the pre-survey and were approved by the hospital’s ethics committee. The ethics approval number was KY2025K394.

#### 1.3.2. Survey Tools

The survey tools consisted of a general information questionnaire for nursing students and the Chinese version of the Peripheral intravenous Catheter Insertion Self-Confidence Scale. The general information questionnaire, determined through literature review and group discussion, included gender, age, education level, number of venipunctures, clinical practice status, theoretical test scores, and practical skills test scores.

#### 1.3.3. Data Collection Methods

The content of the Chinese version of the Peripheral intravenous Catheter Insertion Self-Confidence Scale was entered into the Wenjuanxing software, with a setting that required all questions to be answered before submission. It was also set to allow only one submission per mobile phone. The questionnaires were distributed via a university teacher in Chongqing who sent a QR code to a class group, or by on-site QR code scanning for nursing students in clinical practice in a tertiary hospital in Tianjin. After the questionnaires were completed, the research team carefully reviewed them to ensure they were complete and valid and to check for incorrectly selected options. Two weeks later, 50 nursing students were randomly selected to complete the questionnaire again by scanning the QR code.During this period, there were no unexpected incidents, changes in the teaching environment, or training intervention events.

#### 1.3.4. Analysis Methods of the Reliability and Validity Test

##### 1.3.4.1. Item Analysis

(1) Critical Ratio Method(LI,Z. et al, 2023): The difference in mean scores between the high-scoring group (top 27%) and the low-scoring group (bottom 27%) for each item was calculated. An independent samples t-test was used to verify if the difference was significant. If the significance level P < 0.05, or the critical ratio (CR) > 3.0, the item’s discriminative power was considered significant and thereby retained. (2) Correlation Coefficient Method(LI,Z. et al, 2023): The correlation coefficient between the total scale score and each item score was calculated to assess their relationship. Items with an absolute correlation coefficient of at least 0.4 and a P-value of less than 0.05 after significance testing were retained, and others were eliminated.

##### 1.3.4.2. Validity Testing

Validity was assessed using content validity and construct validity. (1) Content Validity: The Delphi method(Neupane & Bhattarai, 2024) was used for expert consultation to evaluate content validity. Experts rated the relevance and representativeness of the scale’s items on a scale from 0 to 4, with 0 being “not relevant” and 4 being “strongly relevant”. An item-level content validity index (I-CVI) of at least 0.78, an average scale-level content validity index (S-CVI/Ave) of at least 0.90, and a universal agreement content validity index (S-CVI/UA) of at least 0.8 indicate good content validity(ZHANG,C. & ZHOU,Y.X., 2020), suggesting that the scale comprehensively and accurately measures the intended concept. (2) Construct Validity: Construct validity tests the consistency between the measurement results and the theoretical framework. Before conducting factor analysis, the preconditions of the Kaiser-Meyer-Olkin (KMO) value and Bartlett’s Test of Sphericity must be met. A KMO value ≥ 0.8 and a Bartlett’s test P < 0.05 are considered suitable for exploratory factor analysis. Exploratory factor analysis used the principal component analysis method and the varimax rotation method. Dimensions were extracted based on an eigenvalue > 1, and items with a factor loading < 0.4 or high loadings on multiple factors were considered for deletion(JIA,X.J. & SHI,B.X., 2024). Confirmatory factor analysis used the maximum likelihood method to evaluate model fit. The criteria for judging the model were: an absolute fit index of chi-square/degree of freedom (χ2/df) < 3.000, a root mean square error of approximation (RMSEA) < 0.080, and relative fit indices of a Normed Fit Index (NFI), Relative Fit Index (RFI), Incremental Fit Index (IFI), Tucker-Lewis Index (TLI), and Comparative Fit Index (CFI) all > 0.900, which indicates a good fit of the structural equation model(YANG,E.M. et al, 2024).

##### 1.3.4.3. Reliability Testing

Reliability was tested using three methods. ① Internal Consistency Reliability: The most commonly used indicator is Cronbach’s α coefficient, which ranges from 0 to 1. A higher α value indicates better internal consistency. An α value of 0.80 or higher indicates good reliability(JIANG,Y. et al, 2005). ② Split-Half Reliability: The split-half reliability was calculated using the Spearman-Brown coefficient. A value greater than 0.70 indicates good split-half reliability(WANG,J.W. et al, 2025). ③ Test-Retest Reliability: Two weeks later, 50 undergraduate nursing students were randomly selected from the original participants to complete the questionnaire again. The Pearson correlation coefficient was used to test the correlation between the two sets of scores. A correlation coefficient > 0.75 indicates good test-retest reliability(LI,C. & XIN,L., 2008).

### 1.4. Statistical Methods

The results obtained from Wenjuanxing were downloaded into an Excel spreadsheet. SPSS 27.0 and AMOS 29.0 software were used for data processing. Following statistical norms, quantitative data that met the normal distribution conditions were described using mean and standard deviation. Count data were presented using frequency statistics and percentages. The significance level was set at α=0.05.

## Results

### 2.1. Results of Cultural Adaptation and Pre-survey

Based on cultural adaptation and pre-survey results, the revised content and reasons are shown in Table 1. The pre-survey participants were aged 19-21, with four males and 16 females, and completion time between 1 and 4 minutes.

**Table 1.**
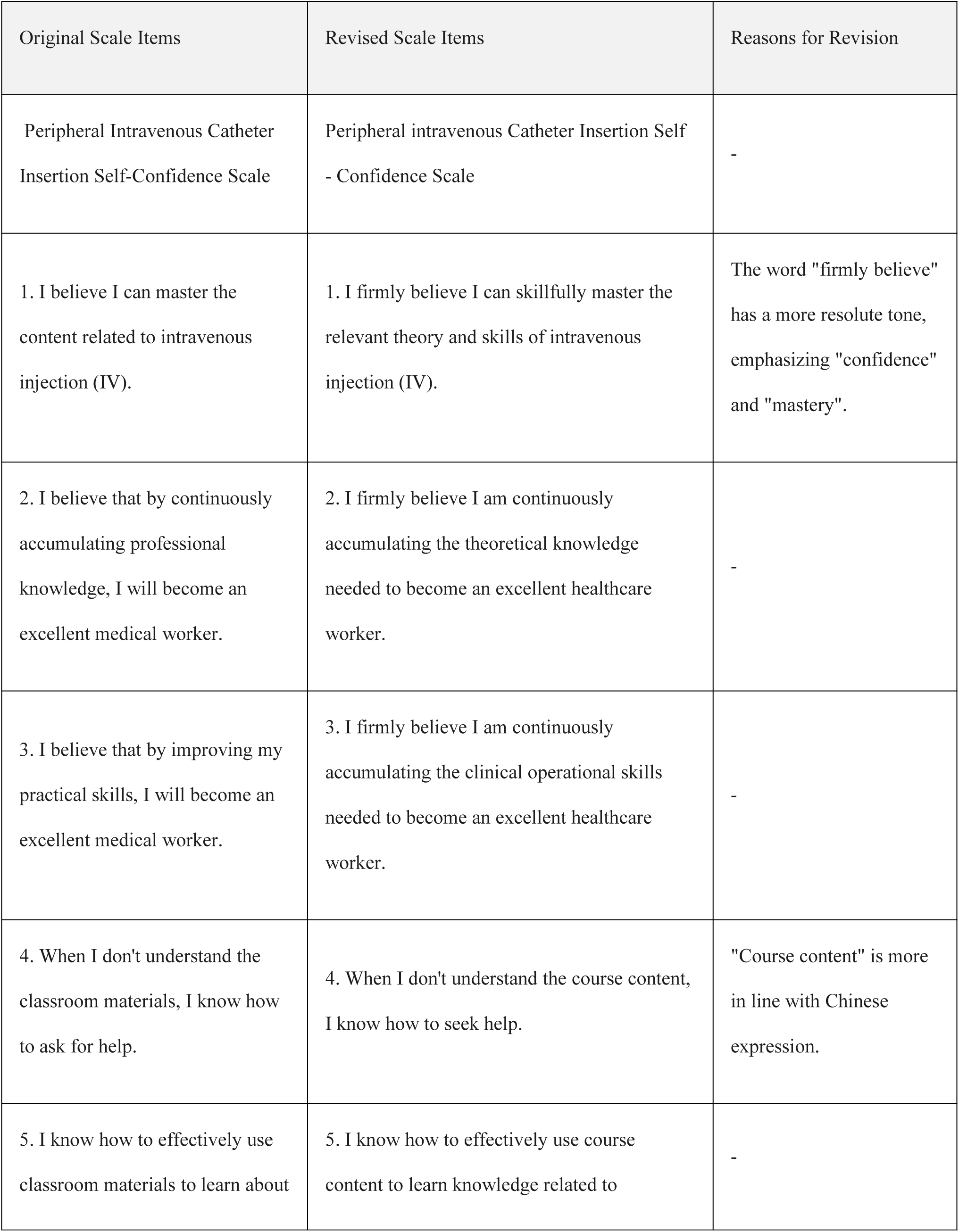
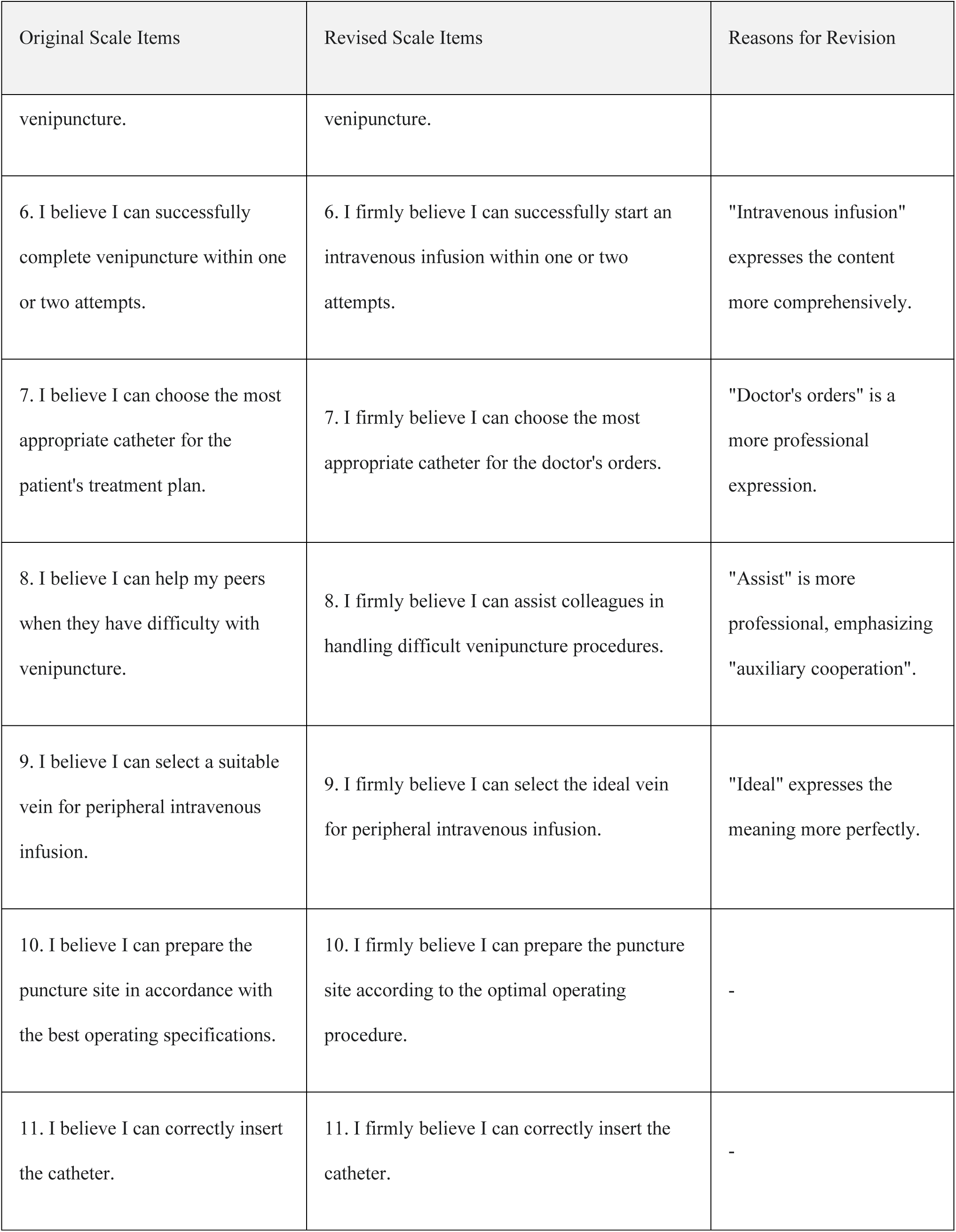
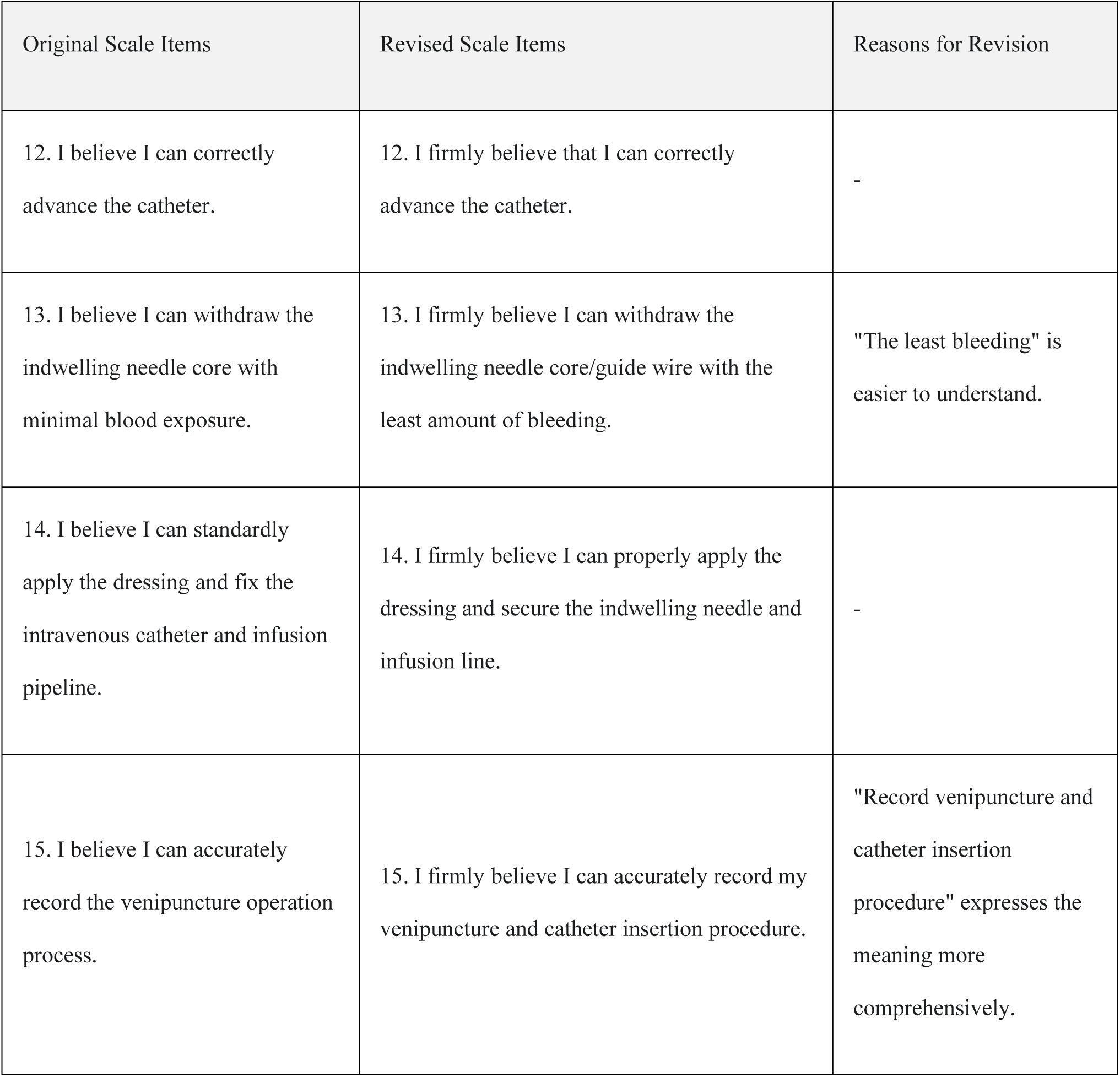
Content and Reasons for Scale Revision.

| Original Scale Items | Revised Scale Items | Reasons for Revision |
| --- | --- | --- |
| Peripheral Intravenous Catheter Insertion Self-Confidence Scale | Peripheral intravenous Catheter Insertion Self - Confidence Scale | - |
| 1. I believe I can master the content related to intravenous injection (IV). | 1. I firmly believe I can skillfully master the relevant theory and skills of intravenous injection (IV). | The word "firmly believe" has a more resolute tone, emphasizing "confidence" and "mastery". |
| 2. I believe that by continuously accumulating professional knowledge, I will become an excellent medical worker. | 2. I firmly believe I am continuously accumulating the theoretical knowledge needed to become an excellent healthcare worker. | - |
| 3. I believe that by improving my practical skills, I will become an excellent medical worker. | 3. I firmly believe I am continuously accumulating the clinical operational skills needed to become an excellent healthcare worker. | - |
| 4. When I don't understand the classroom materials, I know how to ask for help. | 4. When I don't understand the course content, I know how to seek help. | "Course content" is more in line with Chinese expression. |
| 5. I know how to effectively use classroom materials to learn about | 5. I know how to effectively use course content to learn knowledge related to | - |
| venipuncture. | venipuncture. |  |
| 6. I believe I can successfully complete venipuncture within one or two attempts. | 6. I firmly believe I can successfully start an intravenous infusion within one or two attempts. | "Intravenous infusion" expresses the content more comprehensively. |
| 7. I believe I can choose the most appropriate catheter for the patient's treatment plan. | 7. I firmly believe I can choose the most appropriate catheter for the doctor's orders. | "Doctor's orders" is a more professional expression. |
| 8. I believe I can help my peers when they have difficulty with venipuncture. | 8. I firmly believe I can assist colleagues in handling difficult venipuncture procedures. | "Assist" is more professional, emphasizing "auxiliary cooperation". |
| 9. I believe I can select a suitable vein for peripheral intravenous infusion. | 9. I firmly believe I can select the ideal vein for peripheral intravenous infusion. | "Ideal" expresses the meaning more perfectly. |
| 10. I believe I can prepare the puncture site in accordance with the best operating specifications. | 10. I firmly believe I can prepare the puncture site according to the optimal operating procedure. | - |
| 11. I believe I can correctly insert the catheter. | 11. I firmly believe I can correctly insert the catheter. | - |
| 12. I believe I can correctly advance the catheter. | 12. I firmly believe that I can correctly advance the catheter. | - |
| 13. I believe I can withdraw the indwelling needle core with minimal blood exposure. | 13. I firmly believe I can withdraw the indwelling needle core/guide wire with the least amount of bleeding. | "The least bleeding" is easier to understand. |
| 14. I believe I can standardly apply the dressing and fix the intravenous catheter and infusion pipeline. | 14. I firmly believe I can properly apply the dressing and secure the indwelling needle and infusion line. | - |
| 15. I believe I can accurately record the venipuncture operation process. | 15. I firmly believe I can accurately record my venipuncture and catheter insertion procedure. | "Record venipuncture and catheter insertion procedure" expresses the meaning more comprehensively. |

### 2.2. General Information of the Participants

A total of 205 nursing students participated in the questionnaire survey, and all were valid, with a 100% response and recovery rate. The results of the general information are detailed in Table 2.

**Table 2.** General information of the nursing students *(n* = 205)

| Item |  | Number | Percentage | Item |  | Number | Percentage |
| --- | --- | --- | --- | --- | --- | --- | --- |
| Gender | Male | 36 | 17.6% | Clinical Practice Status | Did not observe | 103 | 50.2% |
|  | Female | 169 | 82.4% |  | Observed but not practiced | 36 | 17.6% |
| Age | 18-20 | 169 | 82.4% |  | Practiced | 66 | 32.2% |
|  | 21-23 | 36 | 17.6% | Theoretical Test Scores | Poor | 4 | 2.0% |
| Education Level | Associate Degree | 200 | 97.6% |  | Average | 128 | 62.4% |
|  | Bachelor's Degree | 5 | 2.4% |  | Excellent | 73 | 35.6% |
| Number of Venipunctures | Less than 3 times | 113 | 55.1% | Practical Skills Test Scores | Poor | 15 | 7.3% |
|  | 3-10 times | 33 | 16.1% |  | Average | 103 | 50.2% |
|  | More than 10 times | 59 | 28.8% |  | Excellent | 87 | 42.4% |

### 2.3. Item Analysis Results

The CR values for each item ranged from 20.974 to 58.084, all significantly higher than 3, with a significance level of P < 0.001, indicating good discrimination for each item. The correlation coefficients between each item score and the total scale score ranged from 0.716 to 0.941, all greater than 0.4, with a significance level of P < 0.001. No items were eliminated.

### 2.4. Validity Test Results

#### 2.4.1. Content Validity

The I-CVI of the scale was 0.875-1.000, the S-CVI/UA was 0.866, and the S-CVI/Ave was 0.966. This indicates good content validity, and each item can effectively reflect the level of self-confidence in peripheral intravenous catheter insertion.

#### 2.4.2. Construct Validity

The KMO value was 0.951, and the chi-square for Bartlett’s test was 5125.413, with a P-value less than 0.001. All indicators showed that the scale’s data structure met the prerequisites for factor analysis and could effectively extract common factors. Based on an eigenvalue > 1, exploratory factor analysis extracted two common factors, which, combined with the scree plot results, led to the final determination of two factors. The cumulative variance explained was 85.839%, indicating a strong explanatory power for the original variables. Based on the content of the items, Factor 1 was named “Theoretical Knowledge” and Factor 2 was named “Operational Skills”.Items 4 and 5 had cross-loadings but were retained for content relevance. The rotated component matrix is shown in Table 3. The fitting indexes of the structural equation model indicated that the initial model had a poor fitting effect. After adding residual path correction through the modification index (MI), the results of each fitting index were as follows: χ²/df=2.22, NFI=0.967, RFI=0.956, IFI=0.981, TLI=0.975, CFI=0.981, and RMSEA=0.077, suggesting that the model fits well. The modified structural equation model is shown in Figure 1

**Table 3.** Rotated component matrix of the Chinese version of the Peripheral Intravenous Catheter Insertion Self-Confidence Scale (*n* = 205)

| Item | Operational Skills | Theoretical Knowledge |
| --- | --- | --- |
| 9. I firmly believe I can select the ideal vein for peripheral intravenous infusion. | 0.908 |  |
| 11. I firmly believe I can correctly insert the catheter. | 0.895 |  |
| 12. I firmly believe I can correctly advance the catheter. | 0.893 |  |
| 8. I firmly believe I can assist colleagues in handling difficult venipuncture procedures. | 0.871 |  |
| 10. I firmly believe I can prepare the puncture site according to the optimal operating procedure. | 0.856 |  |
| 13. I firmly believe I can withdraw the indwelling needle core/guide wire with the least amount of bleeding. | 0.852 |  |
| 7. I firmly believe I can choose the most appropriate catheter for the doctor's orders. | 0.825 |  |
| 15. I firmly believe I can accurately record my venipuncture and catheter insertion procedure. | 0.805 |  |
| 14. I firmly believe I can properly apply the dressing and secure the | 0.797 |  |
| indwelling needle and infusion line. |  |  |
| 6. I firmly believe I can successfully start an intravenous infusion within one or two attempts. | 0.738 |  |
| 3. I firmly believe I am continuously accumulating the clinical operational skills needed to become an excellent healthcare worker. |  | 0.868 |
| 2. I firmly believe I am continuously accumulating the theoretical knowledge needed to become an excellent healthcare worker. |  | 0.817 |
| 1. I firmly believe I can skillfully master the relevant theory and skills of intravenous injection (IV). |  | 0.807 |
| 5. I know how to effectively use course content to learn knowledge related to venipuncture. | 0.560 | 0.719 |
| 4. When I don't understand the course content, I know how to seek help. | 0.507 | 0.707 |
| Eigenvalue | 11.770 | 1.105 |
| Cumulative Variance Explained (%) | 53.901 | 85.839 |

**Figure 1.**
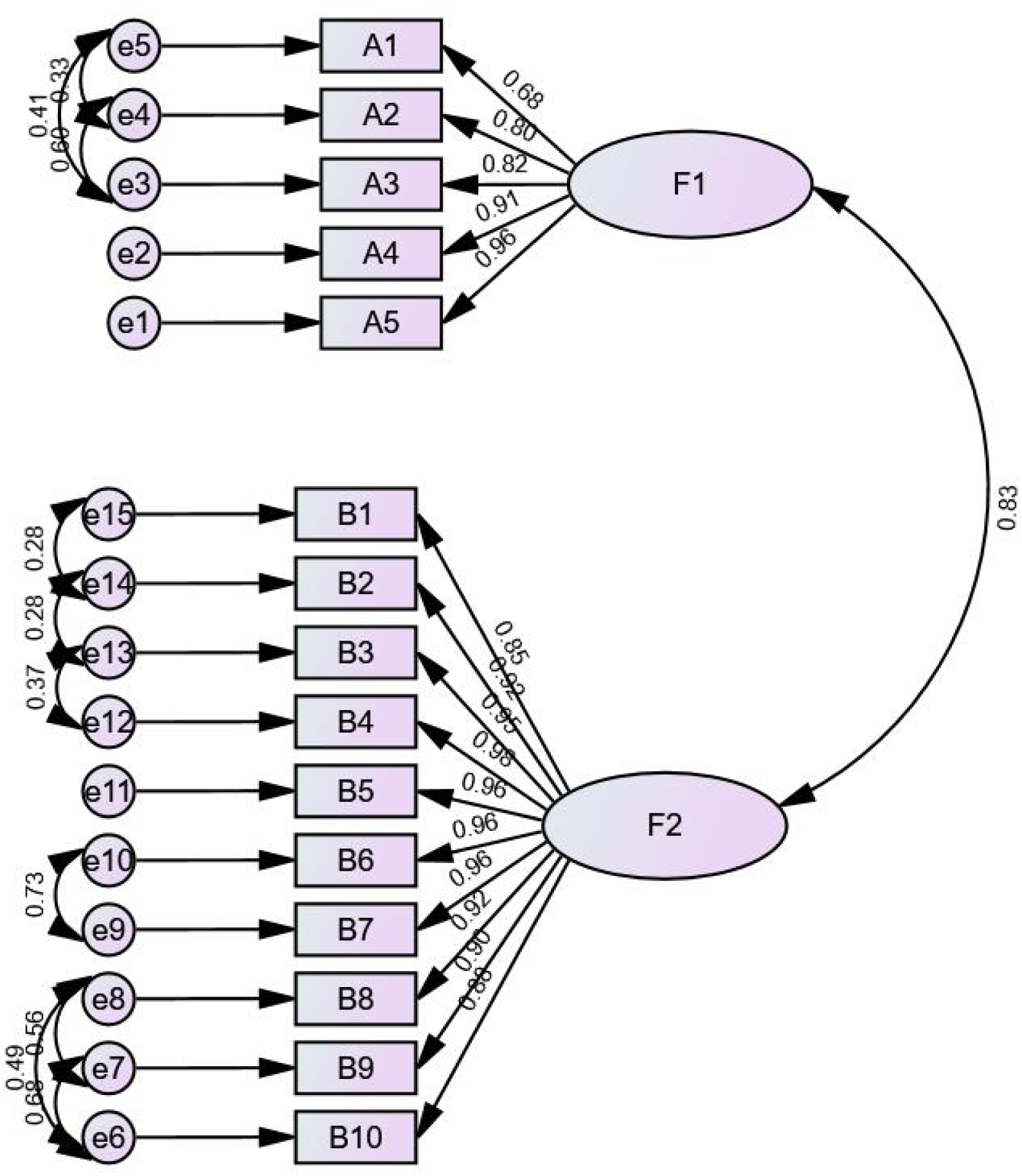
Modified Structural Equation Model Diagram Note. F1=Theoretical Knowledge. F2=Operational Skills

### 2.5. Reliability Test Results

The reliability test results for the Chinese version of the Peripheral intravenous Catheter Insertion Self-Confidence Scale were good. Internal consistency analysis showed that the total Cronbach’s α coefficient was 0.979. The Cronbach’s α coefficients for the “theoretical knowledge” and “operational skills” dimensions were 0.935 and 0.985, respectively. In the split-half reliability test, the total split-half reliability was 0.944. The split-half reliability coefficients for the two dimensions were 0.891 and 0.987. The test-retest reliability results showed a total test-retest correlation coefficient of 0.985. The test-retest reliabilities for the two dimensions were 0.984 and 0.973, respectively. All reliability indicators were at a high level.

## Discussion

### 3.1. The Chinese Version of the Peripheral intravenous Catheter Insertion Self-Confidence Scale Has Good Reliability

Reliability is used to evaluate the consistency of measurement results from multiple measurements or under different conditions. A higher scale reliability indicates a more reliable and stable measurement tool(YE,W.Q. et al, 2023). The C-PVCS shows high reliability, with Cronbach’s α >0.93, split-half >0.89, and test-retest >0.97, indicating consistency and stability.

### 3.2. The Chinese Version of the Peripheral intravenous Catheter Insertion Self-Confidence Scale Has Good Validity

Validity refers to the degree to which a research tool reflects the content it is intended to study. A higher validity indicates that the scale more accurately reflects the studied content(DU,R.F. et al, 2020). The content validity study results showed that the scale’s I-CVI and S-CVI metrics exceed thresholds, indicating that the scale content can accurately reflect the level of self-confidence in peripheral intravenous catheter insertion. Construct validity reflects the scale’s internal factor structure. The exploratory analysis extract two common factors,compared to the original scale, after localization, combined the “preparation and securement self-confidence” and “insertion self-confidence” into a single “operational skills” dimension. Item 1 of the original scale was attributed to the “theoretical knowledge” dimension. This may be related to the cultural differences between China and the West and differences in teaching curriculum design. Catheter preparation and insertion both belong to the operational skills aspect, which is more consistent with the educational model for nursing students in China.Items 4 and 5 have high factor loadings on both dimensions. However, after repeated discussions by the expert group, it is considered that these two items are irreplaceable for measuring nursing students’ confidence in theoretical knowledge from the perspective of content validity, so it was decided to retain them

### 3.3. The Chinese Version of the Peripheral intravenous Catheter Insertion Self-Confidence Scale Has Good Application Value

Peripheral intravenous catheter insertion is one of the most basic invasive operational skills in clinical practice, and it is a fundamental skill that nursing staff must master. Mastering this skill is crucial for improving clinical practice and patient safety(García-Expósito et al, 2021). The Peripheral intravenous Catheter Insertion Self-Confidence Scale can effectively identify nursing students with poor self-confidence, allowing for targeted training and interventions to improve their confidence and success rate.This scale can serve as a teaching assessment tool, guiding teaching staff to adopt targeted teaching interventions based on the score differences of nursing students in the two dimensions of “theoretical knowledge” and “operational skills”. For nursing students with insufficient confidence in theoretical knowledge, theoretical knowledge competitions or case analyses can be added; for those with insufficient confidence in operational skills, simulated practical training such as virtual simulation or OSCE (Objective Structured Clinical Examination) can be provided to systematically improve their confidence and skill levels. The C-PVCS addresses a gap in China, where prior studies used unvalidated tools(CHEN,M. R. & ZHANG,X., 2024; PENG,N. et al, 2020). Its items are specific and highly targeted, consists of multiple-choice questions, is easy to complete, takes 1-4 minutes to fill out, and is highly operable. it is practical for nursing education.

## Conclusion

This study obtained the Chinese version of the Peripheral intravenous Catheter Insertion Self-Confidence Scale through translation, cultural adaptation, and a pre-survey, and tested its reliability and validity among nursing students. The Chinese version of the scale has good reliability and validity and can be used to evaluate the self-confidence level of nursing students in peripheral intravenous catheter insertion. This study uses convenience sampling, and the research subjects are mainly concentrated in specialist nursing students, and the sample size of undergraduate and graduate nursing students is small, which may affect the generality of the results. Future studies may consider expanding sample sizes and employing methods such as stratified sampling to improve sample representativeness. In addition, while we have retained some of the entries with double loading through expert opinion, this also suggests the need to further validate their structural validity in a wider sample in the future. The study subjects were nursing students, but future research could include newly hired nurses to explore its applicability and differences in self-confidence among that group.

## Data Availability

All data produced in the present study are available upon reasonable request to the authors

## Acknowledgements

We thank all nursing trainees for completing the surveys and their mentors for organising data collection. We also thank all colleagues at the Department of Nephrology, The Second Hospital of Tianjin Medical University, for helpful comments.

## References

Chen, M. R. & Zhang, X. (2024). Application effect of virtual simulation technology in venipuncture for nursing interns. Fujian Medical Journal, 46(1), 130~133. 10.20148/j.fmj.2024.01.036

Cruz, J. P., Baigulina, B., Shalkenova, Z., Tau, G., Dossymbayeva, E., & Kostauletova, A. (2023). Investigating the Kazakhstani Pediatric Nurses’ intraintravenous catheter management knowledge and confidence: A cross-sectional study. Nurse Education in Practice, 73, 103816. 10.1016/j.nepr.2023.103816

Du, R.F., Wang, P.P., Chen, C.Y., & Wang, T. (2020). Chinesization of Cardiac Patients Learning Needs Inventory(CPLNI) and its reliability and validity test. CHINESE NURSING RESEARCH, 34(11), 1909~1914. 10.12102/j.issn.1009-6493.2020.11.008

Engum, S. A., Jeffries, P., & Fisher, L. (2003). Intraintravenous catheter training system: Computer-based education versus traditional learning methods. The American Journal of Surgery, 186(1), 67~74. 10.1016/S0002-9610(03)00109-0

García-Expósito, J., Reguant, M., Canet-Vélez, O., Ruiz Mata, F., Botigué, T., & Roca, J. (2021). Evidence of learning on the insertion and care of peripheral intravenous catheters in nursing students: A mixed study. Nurse Education Today, 107, 105157. 10.1016/j.nedt.2021.105157

Guo, J.Y. & Li, Z. (2012). The process and evaluation criteria for scale introduction. Chinese Journal of Nursing, 47(3), 283~285. 10.3761, j.issn.0254—1769.2012.03.039

Jacobs, L. (2022). Peripheral Intraintravenous Catheter Insertion Competence and Confidence in Medical/ Surgical Nurses. 10.1097/NAN.0000000000000487

Jiang, Y., Shen, N., & Zhou, S, F. (2005). Methods for developing scales and assessing their psychometric properties in nursing research. Chinese Journal of Nursing Education, 4, 174~176.

Jia, X.J. & Shi, B.X. (2024). Peritoneal Dialysis Care Burden Questionnaire:translation and validation. Chinese Journal of Nursing, 59(21), 2633~2639. 10.3761/j.issn.0254—1769.2024.21.011

Keleekai, N. L., Schuster, C. A., Murray, C. L., King, M. A., Stahl, B. R., Labrozzi, L. J., Gallucci, S., LeClair, M. W., & Glover, K. R. (2016). Improving Nurses’ Peripheral Intraintravenous Catheter Insertion Knowledge, Confidence, and Skills Using a Simulation-Based Blended Learning Program: A Randomized Trial. Simulation in Healthcare: The Journal of the Society for Simulation in Healthcare, 11(6), 376~384. 10.1097/SIH.0000000000000186

Li, C. & Xin, L. (2008). Research on evaluation methods for reliability and validity of questionnaires. Chinese Journal of Health Statistics, 5, 541~544.

Li, Z., Zhou, M., Chen, L.N., Qu, R.J., Chen, S.N., & Wang, H.L. (2023). Sinicization of the quality of working life questionnaire for cancer survivors and the test of its renability and validity. Chinese Journal of Nursing, 58(6), 695~700. 10.3761/j.issn.0254—1769.2023.06.008

Li, L., Ma, C.Y., Liu, Y., Tang, M.L., & Luo, Y.L. (2024). A Comparative Analysis of the Practice Status before and after the Revision of the“Nursing Practice Standards for Intraintravenous Therapy“. Chinese Health Quality Management, 31(8), 53~57, 68. 10.13912/j.cnki.chqm.2024.31.8.11

Marchionni, C., Lavigne, G., & Connolly, M. (2024). Validation of the Nursing Student Peripheral Intraintravenous Catheter Insertion Self-Confidence Scale. Journal of Nursing Measurement, 32(3), 382~390. 10.1891/JNM-2022-0082

Marsh, N., Larsen, E. N., Takashima, M., Kleidon, T., Keogh, S., Ullman, A. J., Mihala, G., Chopra, V., & Rickard, C. M. (2021). Peripheral intraintravenous catheter failure: A secondary analysis of risks from 11, 830 catheters. International Journal of Nursing Studies, 124, 104095. 10.1016/j.ijnurstu.2021.104095

Neupane, S. M., & Bhattarai, P. C. (2024). Constructing the scale to measure entrepreneurial traits by using the modified delphi method. Heliyon, 10(7), e28410. 10.1016/j.heliyon.2024.e28410

Özbay, H., Su, S., Durgun, H., & Marchionni, C. (2025). Validity and Reliability of Turkish Version of the Nursing Student Peripheral Intraintravenous Catheter Insertion Self-Confidence Scale: Methodological Study. Turkiye Klinikleri Journal of Nursing Sciences, 17(1), 184~194. 10.5336/nurses.2024-105202

Peng, N., Zhang, Q., Qian, Y., Song, J.F., Yang, Z.H., & Liu, L. (2020). Application of venipuncture virtual simulation system in intravenous indwelling needle training for nurses who accepted normalized training. Nursing Practice and Research, 17(16), 138~141. 10.3969/j.issn.1672-9676.2020.16.053

Schuster, C., Stahl, B., Murray, C., & Glover, K. (2016). Development and Testing of an Instrument to Measure Short Peripheral Catheter Insertion Confidence. Journal of Infusion Nursing, 39(3), 159~165. 10.1097/NAN.0000000000000166

Wang, J.W., Wang, M., Gu, H.X., Xu, R., An, W.H., & Yi, Q.F. (2025). Translation and validation of the Sense of Belonging in Nursing School Scale. Journal of Nursing Science, 40(5), 84~88. 10.3870/j.issn.1001-4152.2025.05.084

Wei, Y.T., Tian, S.M., Yang, J., Yu, L.H., Ni, F., Fan, Y.Q., Xiao, Y., Xi, Z.Y., Sha, J.Y.& Liu, C. (2025). Sinicization of Evidence-Informed Decision-Making Competence Measure for nurses and its reliability and validity tes. Chinese Journal of Nursing, 60(6), 736~742. 10.3761/j.issn.0254-1769.2025.06.015

Yang, E.M., Lin, X.Q., Wang, Z., & Yuan, S.S. (2024). Structural Equation Modeling Analysis of the Influence Mechanism of Professional Innovation Performance in Public Hospitals. Chinese Health Economics, 43(3), 44~48. https://kns.cnki.net/KCMS/detail/detail.aspx?dbcode=CJFQ&dbname=CJFDLAST2024&filename=WEIJ202403010

Ye, W.Q., Gao, G.Z., Li, H.Y., Feng, P.Y., & Ji, M.J. (2023). Chineseization and reliability and validity test of Pulmonary Rehabilitation Adapted Index of Self⁃Efficacy. Chinese Nursing Research, 37(2), 216~220. 10.12102/j.issn.1009-6493.2023.02.006

Zhang, C. & Zhou, Y.X. (2020). Analysis on errors regarding content validity index used in evaluation of measurement tools in Chinese nursing literature. Journal of Nursing Science, 35(4), 86~88, 92. 10.3870/j.issn.1001—4152.2020.04.086

